# Online psychometric performance of the Athens Insomnia Scale among Colombian people

**DOI:** 10.1101/2024.02.28.24303466

**Authors:** Adalberto Campo-Arias, Carmen Cecilia Caballero-Domínguez, John Carlos Pedrozo-Pupo

## Abstract

**Objective:** The study aimed to perform confirmatory factor, internal consistency, and differential item functioning analyses of AIS in two Colombian online samples.

**Methods:** Two samples were recruited online at different times. A secondary validation study was conducted with the participation of adults. The first sample was adults from the general population (n = 700, between 18 and 76 years, M = 37.05, SD = 12.70, and 68% were female adults). The second sample included patients with asthma or chronic obstructive pulmonary disease (n = 292, between 18 and 96 years (M = 60.44, SD = 17.57, and 64.73% were female). Separated confirmatory factor analyses and Cronbach’s alpha and McDonald’s omega were computed for each factor. Moreover, gender DIF was calculated using Kendall’s tau.

**Results:** The data adjusted better for the AIS two-dimensional model in the general population sample (chi-squared of 111.25 [df of 19, p < .001], RMSEA of .08, CFI of .96, TLI of .95, and SRMR of .04) and the clinical sample (chi-squared of 96.83 [df of 19, p < .001], RMSEA of .12, CFI of .94, TLI of .91, and SRMR of .05). The internal consistency was high, both Cronbach alpha and McDonald’s omega of .88 in the general population sample, and Cronbach’s alpha of .85 and McDonald’s omega of .88 in the clinical sample. Gender non-differential item functioning was not found among any sample.

**Conclusions:** AIS shows a two-dimensional structure, high internal consistency, and gender non-differential item functioning in two online samples in Colombia. It is necessary to corroborate these findings in other samples.

---

Over the last decade, studies based on Internet samples have grown exponentially with the coronavirus pandemic (De Boni, 2020). However, samples obtained by electronic surveys are criticized for sampling problems, possible unclear answering instructions, or low response rates (Comley, 2002; Evans & Mathur, 2018). Sampling is not a limitation for validation studies because internal validity is sufficient for these investigations. After all, sample representativeness is crucial in the stage of construction of the content validity of an instrument. Assessing a device’s psychometric performance with some trajectory, such as the Athens Insomnia Scale (AIS), among new samples is more relevant than the sample size (Streiner et al., 2015).

Validation studies should be repeated repeatedly in populations and samples with different backgrounds because the performance of these instruments can present significant variations between samples (Campo-Arias & Pineda-Roa, 2022). The consistent findings in various studies confirm the usefulness of screening tools for a specific purpose (Bair & Blais, 2010).

The AIS is one of the most used instruments to screen insomnia problems in clinical and general populations (Soldatos et al., 2000). Through personal or direct application, the AIS has consistently shown high internal consistency, between .73 and .93, in different languages and communities (Campo-Arias et al., 2020; Fornal-Pawłowska et al., 2011; Gómez-Benito et al., 2011; Heidari et al., 2022; Jeong et al., 2015; Nenclares & Jiménez-Genchi, 2005; Niu et al., 2023; Okajima et al., 2013; Soldatos et al., 2000; Sun et al., 2011). However, the dimensional structure can vary from one to three dimensions, according to the participants (Baize et al., 2023; Campo-Arias et al., 2020; Fornal-Pawłowska et al., 2011; Gómez-Benito et al., 2011; Heidari et al., 2022; Jeong et al., 2015; Manzar et al., 2022; Niu et al., 2023; Okajima et al., 2013; Sirajudeen et al., 2020; Soldatos et al., 2000). Moreover, the AIS recently showed a gender differential item functioning (DIF), except for item 2 (awakenings during the night) among cancer patients (Lin et al., 2020).

Similarly, during the COVID-19 pandemic, through an online application, the global AIS has presented excellent internal consistency among adults from the general population, between .82 and .93 (González-González et al., 2023; Mena & Calderón, 2022; Sattler et al., 2023). The few studies available through online applications show a structure with one or two dimensions. Mena and Calderón (2022) retained one dimension among Salvadorian adults. González-González et al. (2023) reported a two-dimensional structure of the AIS in a sample of Mexican adults. Similarly, Sattler et al. (2023) also observed the same two-dimensional structure in a non-clinical sample of German adults; the first dimension (sleep problems) showed McDonald’s omega of .73 and the second (daytime dysfunction) of .93. Nevertheless, gender DIF of the AIS online application has been unreported (González-González et al., 2023; Mena & Calderón, 2022; Sattler et al., 2023).

It is necessary to be confident of AIS’s psychometric performance through online administration; similar results guarantee the validity and reliability of the instrument since it has shown acceptable performance indicators compared to the clinical interview (Chiu et al., 2016). These results may provide a basis for routine screening for sleep problems in the general population through online surveys and teleconsultation or telemedicine (Almathami et al., 2020).

The study aimed to perform confirmatory factor (CFA), internal consistency, and DIF of the AIS in a Colombian online sample of the general population and other clinical outpatients (asthma and chronic obstructive pulmonary disease patients).

## Method

### Design and sample

A secondary validation study was done. The researchers invited to participate to adults residing in Colombia. The first sample was 700 adults from the general population; they were between 18 and 76 years old (M = 37.05, SD = 12.70), and 68% of participants were women. The second sample was a clinical population of 292 asthma and chronic obstructive pulmonary disease (COPD) outpatients. They were aged between 18 and 96 years (M = 60.44, SD = 17.57), and 64.73% of patients were females.

### The scale

During the Colombian lockdown, participants completed an online questionnaire, which included demographic information and AIS items. AIS is an eight-item instrument based on non-organic insomnia criteria from the International Classification of Mental Disorders. Each item provides four response options rated from zero to three, and consequently, the total scores can be from zero to 24; the higher the score, the higher the risk of insomnia (Soldatos et al., 2000).

### Procedure

Participants in both groups completed an anonymous online questionnaire (name, email, or IP address was not requested) that requested demographic information and included the AIS in an item-by-item format. This format was preferred because the grid format can induce more errors in the response (Revilla et al., 2017; Stern et al., 2016). The link was available to participants in the general population between 30 March and 8 April 2020 and participants with asthma or COPD between 1 April and 31 May 2020.

### Statistical analyses

A CFA was performed to test the dimensionality; one- and two-dimensional structures were tested. It was computed several goodness-of-fit coefficients: Normalized chi-squared, Root Mean Square Error of Approximation (RMSEA) and the 90% confidence interval (90% CI), Comparative Fit Index (CFI), the index Tucker-Lewis (TLI) and Standardized Mean Square Residual (SRMR). In the best conditions, the normalized chi-square was expected to be greater than 3.00 (Hair et al., 2006), RMSEA and SRMR with values around 0.06, and CFI and TLI values higher than 0.89 (Hu & Bentler, 1999). At least three coefficients within the desirable values may be sufficient to accept the analyzed data fit (Hu & Bentler, 1999).

Cronbach’s alpha (1951) and McDonald’s omega (1970) were computed as indicators of internal consistency; values greater than .70 were taken as acceptable (Streiner et al., 2015). Kendall’s tau (κ) (1938) was calculated to test gender DIF; κ < .20 was taken as a non-differential response (Hambleton, 2006). The calculations were made with the Jamovi and STATA programs.

### Ethical issues

The research ethics board of the Universidad del Magdalena at Santa Marta, Colombia, reviewed and approved the project in an ordinary meeting (Minute 002 of 26 March 2020). According to Helsinki’s Declaration, participants signed an online informed consent after knowing the research objectives (World Medical Association, 2018).

## Results

### CFA

Poor indicators of goodness of fit were seen for the one-dimensional factor solution in both samples; then, it was rejected. On the other hand, the two-dimensional solution presented better adjustment; in the general population, it got acceptable four out of five indicators of goodness of fit, and in the asthma and COPD samples, three out of five were acceptable. See Table 1.

**Table 1.** Indexes of goodness of fit for the one- and two-dimensional model.

| Index | General population |  | Clinical sample |  |
| --- | --- | --- | --- | --- |
|  | One dimension | Two dimensions | One dimension | Two dimensions |
| $X^2$ (df, $p <$) | 295.91 (20, .001) | 111.25 (19, .001) | 195.14 (20, $< .001$) | 96.83 (19, $< .001$) |
| $X^2 / df$ | 14.79 | 5.86 | 9.71 | 5.10 |
| RMSEA (90% CI) | .12 (.14-.16) | .08 (.06-.10) | .18 (.16-.20) | .12 (.10-.15) |
| CFI | .89 | .96 | .87 | .94 |
| TLI | .85 | .95 | .82 | .91 |
| SRMR | .06 | .04 | .07 | .05 |

### Internal consistency

In the general population sample, global AIS showed Cronbach’s alpha and McDonald’s of .88: Factor I (night sleep) presented Cronbach’s alpha of .85 and McDonald’s omega of .86, and factor II and (daytime dysfunction) Cronbach’s alpha of .80 McDonald’s omega of .81. Furthermore, in asthma and COPD sample, global AIS presented Cronbach’s alpha of .85 and McDonald’s omega of .88, the factor I showed Cronbach’s alpha of .80 and McDonald’s omega of .82, and factor II Cronbach’s alpha of .82 and McDonald’s omega of .83.

### Differential item functioning

There were no differences in functioning according to gender. The general population κ was between .02 and .10, and the asthma and COPD samples were between κ -.02 and .09.

## Discussion

The study shows that the AIS two-dimensional model adjusts to the data and asthma, COPD, and general population samples. Also, AIS presents a high internal consistency and gender non-differential item functioning.

In the present study, the finding of the two-dimensional structure for AIS is consistent with a previous study with direct application, Okajima et al. (2013), in Japan, observed in 477 outpatients with insomnia and 167 people from the community, Heidari et al. (2022), in Iran, with 155 patients; Manzar et al. (2022), in Saudi Arabia among 563 nurses; and Baize et al. (2023), in France, among 296 competitive athletes: The first five items they formed the factor “sleep problems” and the last three, the factor “daytime dysfunction.”

However, the finding of current study differs from other analyzes in which a one-dimensional structure was observed from the original study by Soldatos et al. (2000), in Greece, with 105 primary care patients with insomnia, 100 psychiatric outpatients, 44 psychiatric inpatients and 50 controls; Fornal-Pawłowska et al. (2011), in Poland, 166 patients with primary insomnia and 196 controls with good sleep; Gómez-Benito et al. (2011), in Spain; Jeong et al. (2015), in South Korea, in 221 firefighters and rescuers; and Campo-Arias et al. (2020), in Colombia, in 1,358 climacteric women. Besides, Sirajudeen et al. (2020), in India, with 434 occupational computer users participating, reported the best solution for a three-dimensional structure of the AIS.

Similarly, online applications have reported two-factor solutions: one-dimension was described among 1,479 Salvadorian adults (Mena & Calderón, 2022), and two-dimension reported in 4,047 Mexican adults (González-González et al., 2023) and 24,809 non-clinical adults in Germany (Sattler et al., 2023). These variations in the dimensionality of the instruments are related to the differences in the response patterns of each participating sample. It is frequently mentioned that this is more a property of the response pattern of the participating population rather than of the measurement scale (Streiner et al., 2015).

In the present study, AIS showed high internal consistency. Findings are consistent with previous studies that found consistency between .73 and .93 for the global scale through direct or online application (Campo-Arias et al., 2020; Fornal-Pawłowska et al., 2011; Gómez-Benito et al., 2011; González-González et al., 2023; Heidari et al., 2022; Jeong et al., 2015; Mena & Calderón, 2022; Nenclares & Jiménez-Genchi, 2005; Niu et al., 2023; Okajima et al., 2013; Sattler et al., 2023; Soldatos et al., 2000; Sun et al., 2011). Moreover, the current study’s findings are consistent with previous publications showing that the AIS’s two dimensions presented high internal consistency (Okajima et al., 2013; Sattler et al., 2023). These findings suggest that the AIS performs similarly in internal consistency regardless of the application form. However, high internal consistency does not guarantee dimensionality (Bair & Blais, 2010).

Furthermore, in the present study, it was observed that AIS items showed non-differential functioning according to gender; this is inconsistent with the Yin et al. findings (2020) in Taiwan, in 30 patients with cancer, in whom they observed that the response showed gender bias, except the item 2. Bias item functioning is always unexpected because inducing an error in the measure (Bair & Blais, 2010; Streiner et al., 2015).

### Practical implications of online applications

These findings contribute to the validity and reliability of AIS. The AIS can be considered for detecting insomnia through surveys (Almathami et al., 2020). The online application of the instrument has the advantage of research. It can have many participants, the time for collecting information is reduced, it usually costs less than face-to-face applications, and the answer is more honest (Comley, 2002; Evans & Mathur, 2018). However, the online application has limitations such as the sample’s representativeness, the fact that it excludes participants with limited access to technology, the difficulty in instructing participants, and the low response rate (Campo-Arias et al., 2022; Evans & Mathur, 2018).

### Strengths and limitations of the study

This research has the strength that it compared the performance of the AIS in two different samples of participants, calculated two indicators of internal consistency (Cronbach’s alpha and McDonald’s omega), and explored the gender DIF usually omitted in the analysis of the AIS performance. However, it had the limitation that the application of another insomnia measure was avoided. This would have allowed us to estimate the convergent validity of the AIS. Future studies with online applications should explore the convergent validity of the instrument (Bair & Blais, 2010; Keszei et al., 2010; Streiner et al., 2015).

## Conclusions

The AIS presents a comparable performance in two online surveys in asthma and COPD samples and from the general population, similar to the traditional application. It is crucial to corroborate these findings in other general population samples and patients with different clinical conditions.

## Data Availability

The data that support the findings of this study are available from the corresponding author upon reasonable request.Universidad del Magdalena, Santa Marta, Colombia.

